# Net and gross efficacies; when a single RCT is not enough and multiple RCTs are impracticable

**DOI:** 10.1101/2025.11.19.25340562

**Authors:** Jean-Pierre Boissel, Evgueni Jacob, Arnaud Nativel, Emmanuelle Bechet, Riad Kahoul, Michael Duruisseaux, Solène Granjeon-Noriot, Jim Bosley, François-Henri Boissel, Emmanuel Pham, Marc Hommel, Claudio Monteiro

## Abstract

This work takes place at two interacting levels. One is a reflection on how to define therapeutic efficacy. The other one is a series of comparative simulation experiments confronting two definitions of therapeutic efficacy.

In real-world settings, treatment efficacy is typically estimated through randomized controlled trials (RCTs), yielding what we define as gross efficacy (GE). However, biases introduced by the enrollment process and imperfect results of randomization can affect GE estimates. In contrast, in a model informed drug development (MIDD) perspective, an in silico clinical trial (ISCT) is unbiased. It is conducted via computational simulations using a quantitative systems pharmacology (QSP) model of disease and treatments and a virtual population. The two (or more) compared treatments are given in turn to the same virtual patient, with the same environment, at the same time, in an as close as possible representation of the whole population of interest, resulting in the prediction of the net efficacy (NE). Because inter-individual and inter-occasion differences are removed, the NE prediction contains no external bias.

In order to explore the two paradigms, GE and NE, clinical trials were simulated using a disease model of advanced EGFR-mutated lung adenocarcinoma (aLUAD), a subtype of Non Small Cell Lung Cancer (NSCLC) and two treatment models, one for the investigational treatment, osimertinib in monotherapy, (A), and one for the control treatment, chemotherapy agents cisplatin and pemetrexed (B). The model used in the current simulations was a simplified model of a model that has proven its credibility in prospectively and blindly predicting accurately the results of real world phase 3 clinical trials, after a thorough validation procedure. The simplified model was applied in two different settings to a large virtual population. The first setting was designed to compute the net efficacy of treatment A versus B. The second setting was set-up to mimic 1,000 real-life, parallel group design RCTs, based on standard sampling theory to compare A and B. The number of patients per arm was selected to be equal to or greater than the currently published trials. As opposed to the first approach, the approach mimicking the RCT provides measures of the gross efficacy. A single RCT was randomly drawn to simulate a phase 3 trial which concludes a clinical development program. While the mean hazard ratios of clinical outcomes calculated by the GE approach and the hazard ratio of clinical outcome given by the NE approach are not very different, they could nevertheless lead to quite different assessments of the population benefit of the new treatment compared to a comparator and to different regulatory decisions. Further, some values of GE from the distribution of “observed” GE were rather far from the mean GE and the NE. The convergence of the GE estimate takes more than a couple of RCTs, showing that a single phase 3 trial is not enough. However, the number of trials sufficient for convergence is impracticable. This work also shows that a randomized phase 3 trial of a genuinely effective therapeutic can easily fail to demonstrate a difference between the two groups of treatment due solely to sampling fluctuations, incorrectly leading to the discontinuation of the investigational drug because of lack of chance. These findings suggest that while ISCTs cannot replace real-world RCTs, they provide valuable insights for establishing clinical development strategy, trial design, trial monitoring, and trial results interpretation.

## Introduction

Model-informed drug development (MIDD) was introduced by the FDA in 2013^1,2^. Modeling, as inspired by long experience in pharmacokinetic modeling, was seen as “a powerful approach to increase efficiency in drug development, to impact regulatory outcomes, and to improve drug benefit–risk balance”. Central to this framework is quantitative systems pharmacology (QSP), which represents an important set of tools used for MIDD^3^. This set covers two different areas of expertise, often opposed but in fact complementary. One is based on statistical modelling of observed data on patients or healthy subjects, and the other on mathematical or logical modelling of mechanistic scientific knowledge drawn from scientific literature, the physiological paradigm.

The *statistical paradigm* was initially employed by clinical experimentalists in the context of clinical trials, prior to QSP. It structures a randomized controlled trial (RCT) along with the experimental design principles (the principles of unbiased comparison). In a RCT, the effects of a treatment on a parent population are estimated through the observation of a sample of patients, which is assumed to be randomly drawn from that population^4,5^. This operation is an intellectual construct as the process of constituting the trial sample is not a true random drawing from the population of interest but rather a constrained selection process influenced by the situation of the investigators network in the health system ^6^ and the investigators interpretation of eligibility criteria ^7^. Rather than being pre-existing to the trial sample, the parent population is derived *from* the sample.

The *physiological paradigm* relies on a mathematical model that encapsulates the mechanistic and epidemiological knowledge (biology, physiology, pathophysiology, pharmacology) about the system of interest along with a simulated population mirroring the real population of interest ^8–10^. It is then no longer necessary to use the sampling theory to make predictions derived from the simulation results since a representation of the entire population of interest is included in the in silico trial. This approach is called knowledge-based modeling to highlight the difference with the previous one, which is mainly data-driven. It has been also quoted as qualitative modeling^11^, although it enables quantitative prediction based on the causal relations extracted from the knowledge.

The first paradigm, by its nature and reliance on data collected at discrete time-points, provides an estimate of the current observable efficacy. In this sense, the RCT based estimate of efficacy is a data driven model. In contrast, the second paradigm which aims to reproduce the dynamics of the disease mechanisms and interactions between the drug and the patient’s body, including, if needed, co-morbidities, in theory enables robust predictions of the “true efficacy”.

However, a bridge exists between the two paradigms, where data-driven and knowledge-based models (KBM) are interwoven. For example, in physiology-based pharmacokinetic models^12^ (PBPK), instead of assuming a series of artificial compartments for drug distribution as in PK modeling, the modeler relies on established interactions between the compound and organs such as the liver and lungs and aims at reproducing mathematically these interactions. In addition, the virtual population of a KBM is partly constructed from the data describing the population of interest.

When assessing the efficacy of a drug, the statistical paradigm relies on statistical tests for decision-making based on available observed data. In contrast, in the knowledge-based approach, such statistical tests become irrelevant, as predictive calculations are made on the comprehensive, ideally one-to-one, representation of the real patients. However, appropriate methods need to be defined to estimate the uncertainty associated with the model predictions^13^.

This fundamental difference between the two paradigms generates a novel definition of the efficacy of a therapy, which this article aims to illustrate.

## Gross and net efficacy

#### Box 1: various types of therapy efficacy

The efficacy of a treatment is a polysemous concept, that mutates depending on given properties of the situation it quantifies:

- Considered endpoints: A drug can be efficacious in decreasing blood sugar but not in decreasing mortality linked with high blood glucose.
- Individuals: The efficacy given by a RCT is a group or average efficacy. It differs from the efficacy a doctor thinks about when he says ‘this treatment has been efficacious in this patient’. Indeed, in that case, the object is individual instead of a group. Moreover, the assertion is based on quite biased comparators: the patient status before treatment and the courses of other patients.
- Comparator: Even unbiased RCTs do not lead to the same type of efficacy. For instance, everything being equal, a parallel group designed trial and a cross-over design do not measure the same type of efficacy. So, among others, the former being more biased than the latter, it is supposed to be less close to the “real” efficacy.

Hence, the efficacy of a new drug is difficult to define, besides being difficult to measure.

#### Box 2: net efficacy

A treatment net efficacy (NE) is an unbiased value of efficacy obtained by comparing the new treatment scenarios with the comparator scenario in the same patients, each one being his or her own control, at the same stage of the disease, the same time, with the same environment. It can be measured at the individual or at the group level. In practice it is only available thanks to ISCT.

A randomized clinical trial (RCT) is conducted according to a well-established methodology^14–17^. In real practice, this method is applied through the following process. Eligible patients are successively recruited from any participating sites to constitute the trial sample. Then, they are randomly allocated to the arms, each defined by their treatment scenarios, the course of which will be compared. Once the trial ends, comparisons between arms resort to the statistical random sample paradigm. The sample on which the trial focused is assumed to have been randomly drawn from a parent population defined in theory by the eligibility criteria, in practice by the sample baseline characteristics. According to this paradigm, would a new trial with the same protocol be carried out in every respect similar to the first, the estimated efficacy would likely be more or less different from the first estimate since the patients will not be exactly the same, depending on the variability in the accessible population from which the samples were drawn. In addition, the new parent population will be different from the first one. By repeating the trial many times, one would obtain the distribution of these estimates among which the true value of the efficacy is assumed to reside. The true value of the efficacy, or “real” efficacy is a concept, as discussed in the discussion section. In theory, within an ideal realization of this paradigm, any deviation between any estimate provided by a real life RCT and the actual, real efficacy comes from the sampling process. In practice, there are other causes of deviation from reality, notably those due to the imperfection of the processes and tools used for the implementation of the methodology of the randomized trial, such as verification of eligibility criteria, breaches in masking for random allocation or, above all, the impossibility of establishing absolutely head-to-head identical arms, which is a requirement for unbiased comparisons.

The efficacy estimated through a randomized trial in real-world settings can therefore be termed as gross efficacy (GE). This implies that the figure found is tainted with residual factors that are impossible to identify and quantify, and therefore to eliminate. These factors are partly associated with patient - and context-dependent characteristics that are challenging to measure effectively.

Implicitly, this is why the regulatory authorities advocate for the replication of “positive” phase 3 trials of a given product, which may be though, unethical and is obviously impossible at a large enough scale in practice. Actually, the observation of the distribution of gross efficacy estimates is all the more illusory since if such repetition of trials were carried out, the attached parent population would change with each trial. Consequently, this distribution can only be inferred theoretically within the framework of the statistical paradigm. Even then, it is impossible to account for the effect of unmeasured parameters on the variability of RCT results.

In contrast, a clinical trial conducted within a virtual environment, defined by a knowledge-based mathematical model and a virtual population, offers a markedly different opportunity.

First, the in silico clinical trial (ISCT) is not subjected to the ethical issues and practical limitations associated with patient eligibility checking and follow-up in real-world trials. Further, the ISCT facilitates addressing challenges such as selecting the optimal drug regimen^8^ and managing compliance variability.

Above all, this approach has the great advantage of allowing the computation of so-called net efficacy (NE). This NE is deemed to approximate more closely the real efficacy than gross efficacy because in the ISCT, the causes tainting the gross efficacy do not intervene (unless voluntarily introduced to mimic more closely a real trial):

- Each virtual patient is his or her own control, whatever are the follow-up duration, the endpoints, the number of treatments to be compared. Thus there is no random allocation bias by design.
- The application of the protocol is completely mastered.
- The group of patients on which the trial relies is not a sample but is a whole population of interest taking into account all the diversity of eligible patients and whose size is limited only by the simulation time.
- Variables or parameters that cannot be measured but which could impact efficacy are identifiable and taken into account.
- Further, the simulated experiment also provides the individual benefit distribution and not just an “average” figure that does not correspond to any real patient.

This approach circumvents current issues that make RCTs results and, consequently, drug clinical development a source of dispute. Issues such as subgroup size, comparison power and the multiplicity of statistical tests are no longer unmanageable puzzles^18^. Pragmatic trials have been proposed to address some limitations of traditional RCTs^19,20^. They are difficult to design and carry out correctly. They do not solve these issues. In an ISCT, because of this net efficacy computation, the use of the sampling statistical paradigm is no longer necessary since the estimate is based on the entirety of the population of interest and not on a sample, more or less randomly drawn from the latter. Metaphorically, this virtual population is comparable to the parent population invoked for the application of the statistical paradigm in a real-life clinical trial. Provided that the virtual population is fully representative of the real population of interest^21^, the ISCT result is a prediction of a “true” efficacy. Unlike the parent population, the virtual population is stable across all the in silico trials with the same objective and design. And it is this “parent” population that the trial is based upon, whereas a real trial is run on a sample of the parent population, which, itself, is imagined. Further, we claim the parent population rarely fits with the real population of interest because the participating sites are neither the whole set of organizations caring about the disease of interest, nor are they a random sample of the whole set. An exception may occur in the case of rare diseases: where centralized management can theoretically allow for the enrollment of all subjects meeting the eligibility criteria.

The credibility of the net efficacy prediction and thus the validation of the underlying mechanistic model, remain key challenges. These questions are not really different from those that arise when the gross efficacy value derived from a real-life trial is used for decision-making. They muster the same types of backgrounds that are required to check, on the one hand, the coherence of available knowledge with the effects of the drug of interest observed in the RCTs and, on the other hand, the consistence of the RCTs outputs with additional well codified clinical observations in real life.

### Two different paradigms

The traditional paradigm for real life RCT is rooted in sampling theory^22,23^. It assumes that the trial sample of eligible patients is randomly drawn from an infinite population, called the parent population, the structure of which is reflected by the trial sample. This relationship permits inferring, from the observed behavior of the trial patients under experimental and control treatments, what is likely to occur in the parent population under the same treatments. The result of that is the inevitable use of inferential statistics to extrapolate the observed trial findings to the parent population, and, a step further, to future patients as long as those patients are within the parent population. The profile of patients who are deemed to beneficiate more from the experimental treatment (at the cost of few side effects) defines the treatment target population, a subset of the parent population, sometimes extended thanks to extrapolation from the available factual evidence obtained in the RCT.

Instead, the simulated trial (ISCT) is run on the whole virtual population of eligible patients, assumed, or, better, proven to represent in size and structure the corresponding real population of interest. There is no need to infer, just to compute and read. As a consequence, there is no need for statistical tests and therefore no p-value. Also, there is no need to extrapolate.

Another more subtle point differentiating the two paradigms merits mention. Real-life trials and their analysis rely on the presently available data. The simulated trial works on a timeless material, the scientific knowledge on disease and treatment effect dynamics. Depending on this knowledge’s strength of evidence, these dynamics will be the same in the future. Thus, running a model representing this knowledge allows prediction of some quantity, here a predicted size of efficacy, instead of an observation. Moreover, as new knowledge emerges, it can be integrated into the model, allowing the prediction to be revised accordingly.

### Illustration

In order to compare the two paradigms, and to explore the net and the gross efficacy concepts, clinical trials were simulated using a disease model, two treatment models and a realistic virtual population. The models were knowledge-based (KBM), sometimes referred to as a qualitative approach. The collected and curated knowledge on the disease and the treatment(s) is translated in mathematical equations. Originally, the in silico comparison was made to blindly predict the results of FLAURA2^24^. After their validation^25^, the models were applied to a virtual population in two different settings to compute efficacy. The first setting was designed to compute the net efficacy of a treatment A against a treatment B on the whole virtual population. The second setting, was set-up to mimic a real-life, parallel group design RCT, based on standard sampling theory to compare A and B. Both settings are two-sided in our illustration, meaning that there is no a priori guess on the better treatment nor they are situations of investigational treatment against control. As opposed to the first approach, the mimicked RCT approach provides measures of the gross efficacy, investigated similarly as what would have been analyzed in in vivo clinical trials, along with illustrative p-values. In both cases, the endpoint was time to progression. The clinical outcome of interest used for this illustration is the time to tumor progression (TTP), therefore, in a context of time-to-event analysis, the metric used to assess the treatment efficacy is the hazard ratio (HR) for progression. HR and TTP differences were displayed in such a way that, respectively, shown HRs were less than 1 and differences in TTP more than 0 for the more efficacious treatment.

## Material and method

### The models

The disease model was a model of a sub-type of NSCLC (Non Small Cell Lung Cancer), the Advanced EGFR-mutated lung adenocarcinoma (aLUAD). The two treatment models were, one for treatment A, osimertinib in monotherapy and one for treatment B, a combination of chemotherapy agents cisplatin and pemetrexed. The disease model used in the current simulations was a simplified model of a model that has proven its credibility in prospectively and blindly predicting accurately the results of real world phase 3 clinical trials, after a thorough validation procedure^26^. The models were simplified to speed up simulations, given the anticipated number of runs. There were a total of 2068 ODEs in the original models while only 1003 in the simplified models. Modeling and simulations used the jinkō platform (Novainsilico, Lyon, France) which mixes artificial intelligence (AI) and mathematical solutions, while the data post-processing and generation of graphical outputs were performed with R v4.3.2.

### The virtual population

The size of the virtual population was N = 2,000. Virtual patients were designed based on inclusion and exclusion criteria consistent with the FLAURA2 trial. Inclusion criteria encompassed adults with confirmed EGFR mutations (exon 19 deletions or L858R substitutions) and untreated advanced or metastatic NSCLC. Exclusion criteria ruled out patients with prior systemic anticancer therapy for advanced disease, significant comorbidities, or contraindications to study medications. There were 112 descriptors for each patient of the virtual population, among them only 7 were measurable (93.7% of hidden descriptors or model parameters). Among these 7, 5 were baseline variables for the gross efficacy mimicked RCTs.

### Computation of the net efficacy

Net efficacy is predicted by comparing the results of running the mechanistic disease model on the eligible virtual population in which each virtual patient acts as his or her own control with fully identical baseline characteristics receiving in turn the compared treatments. The settings, the time and the patients being the same, net efficacy value is entirely free of bias. Net efficacy prediction is associated with a Prediction Percentile Interval (PPI). Since there is no standard metric for catching uncertainty of predicted quantity in such a setting, we compute the PPI. It is a measure of the uncertainty of the prediction as if the predicted value would be obtained in real life with varying populations. By construct, it includes both modeling errors (the model uncertainty component) and the inter-patient variability as reflected in the virtual population (the population-linked prediction uncertainty). Here, it is obtained with the 2.5% and 97.5% quantiles of the bootstrapped distribution of the predicted value. In this study, the 95% PPI associated with the metric of interest (e.g. HR) was obtained by calculating this measure for 1,000 (n) populations of 300 patients, a number chosen to be a reasonable proportion of the whole population of interest (2,000 patients). This 95% PPI derived by bootstrapping should not be confused with a 95% CI, the former measuring the degree of uncertainty in the model’s predictions, the later measuring, in a real-life context, the uncertainty linked to sampling.

### Mimicked RCT approach: distribution of gross efficacy estimate over 1,000 replications of the same mimicked RCT

The RCT experimental setting is mimicked by randomly sampling a set of patients (N = 200) from the eligible virtual population. The mimicked trials sample size was computed via the formula extracted from [Schoenfeld DA. Sample-size formula for the proportional-hazards regression model. Biometrics. 1983 Jun;39(2):499-503. PMID: 6354290], based on the following hypotheses: alpha risk = 5%, beta risk = 10%, expected HR = 0.59, recruitment duration = none (given the in silico context), follow-up duration = 24 months, median survival time in control arm = 8,1 months, allocation ratio = 1:1, censoring rate = 20%.

Then each patient of the sample was allocated randomly without replacement to two trial arms (treatment A and treatment B) with a 1:1 ratio, running the clinical trial by applying the models to each virtual patient and analyzing results using the standard statistical approach. 1,000 independent mimicked RCTs comparing treatment A to treatment B were performed to illustrate random variations of the gross efficacy estimate related to sampling. Out of these 1,000 iterations, a single randomly selected case, mimicking a phase 3 trial situation, the main milestone of a clinical development, is also presented using the standard statistical paradigm.

## Results

### Net and gross efficacies

The graphical representation of the cumulative average HR obtained over the 1,000 mimicked RCT replicates as a function of the number of achieved repetitions (Figure 1) shows that gross efficacy estimate converges to a value which can be seen as the “real” gross efficacy (HR^avg^_gross_) of around 0.60 for time to progression (TTP). This “real” gross efficacy is close to the net efficacy on TTP that can be obtained with the in silico approach, HR_net_ = 0.59 (see also Table 1).

**Figure 1.**
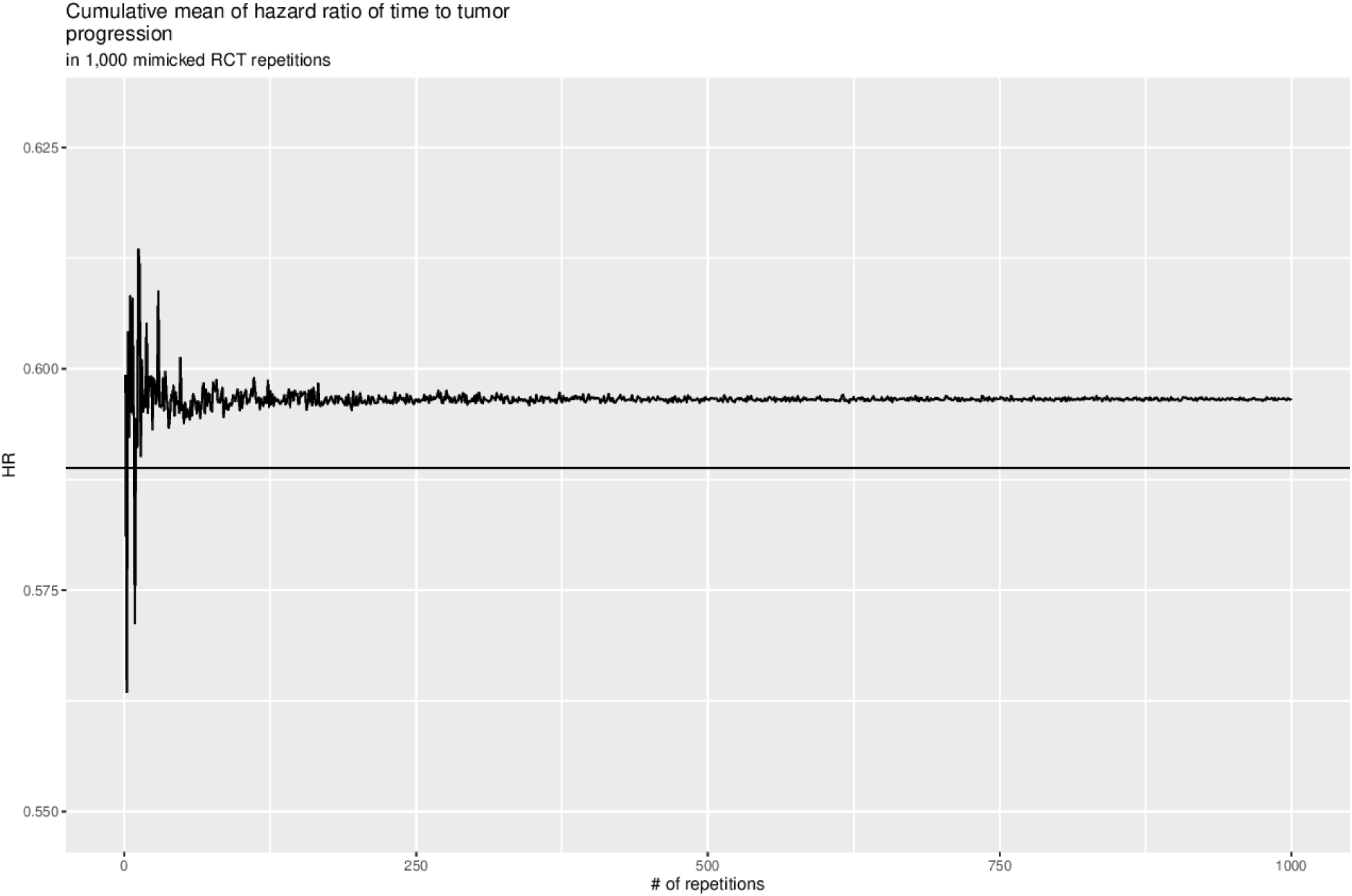
Assessment of the convergence of HR over repetitions of mimicked RCT. Convergence of time to progression HR over 1,000 mimic RCTs is represented via the evolution of the cumulative mean HR as a function of the number of mimic RCT replicates (black line: average value).

**Table 1.** Simulated values of efficacy on the primary outcome (TTP)

|  | Net Efficacy (NE) | Gross Efficacy (GE) |  |  |
| --- | --- | --- | --- | --- |
|  |  | Average | Closest MRCT** | A random Phase 3 MRCT |
| Value | 0.59 | 0.60 | 0.60 | 0.72 |
| Uncertainty marker<sup>°</sup> | [0.55; 0.63] | [0.60, 0.61] | [0.43; 0.83] | [0.52; 0.99] |
| Prediction uncertainty marker<sup>*</sup> | [0.52; 0.67] |  |  |  |
| Number of prevented progressions<sup>°°</sup> | 240 | 235 | 235 | 196 |

Legend: <sup>°</sup> 95% Confidence interval computed on the distribution of GE (average), on the parent population of the mimicked RCTs (MRCT), and PPI on the virtual population (NE); <sup>\*</sup> PPI; <sup>°°</sup> for a treated population of 2,000 patients, assuming a linear multiplicative effect model and that the HRs computed in this work are good estimate of the ratio prevented event rate under treatment B over prevented event rate under treatment A; <sup>\*\*</sup> mimicked RCT which gives a GE value closest to the average GE

The simulated empirical distributions of gross efficacy estimated by mimicked RCT, represented by the distribution of HR between treatment A and treatment B obtained from the 1,000 mimicked RCT replicates are shown on Figure 2. On the same figure, are also shown (i) the gross efficacy estimate obtained with the randomly selected mimicked RCT (blue dashed line) with the standard statistical methodology - see below - and (ii) the net efficacy estimate obtained with the in silico paradigm approach and the entire virtual population (red dashed line). The range of observed HR (HR_gross_) is rather large.

**Figure 2.**
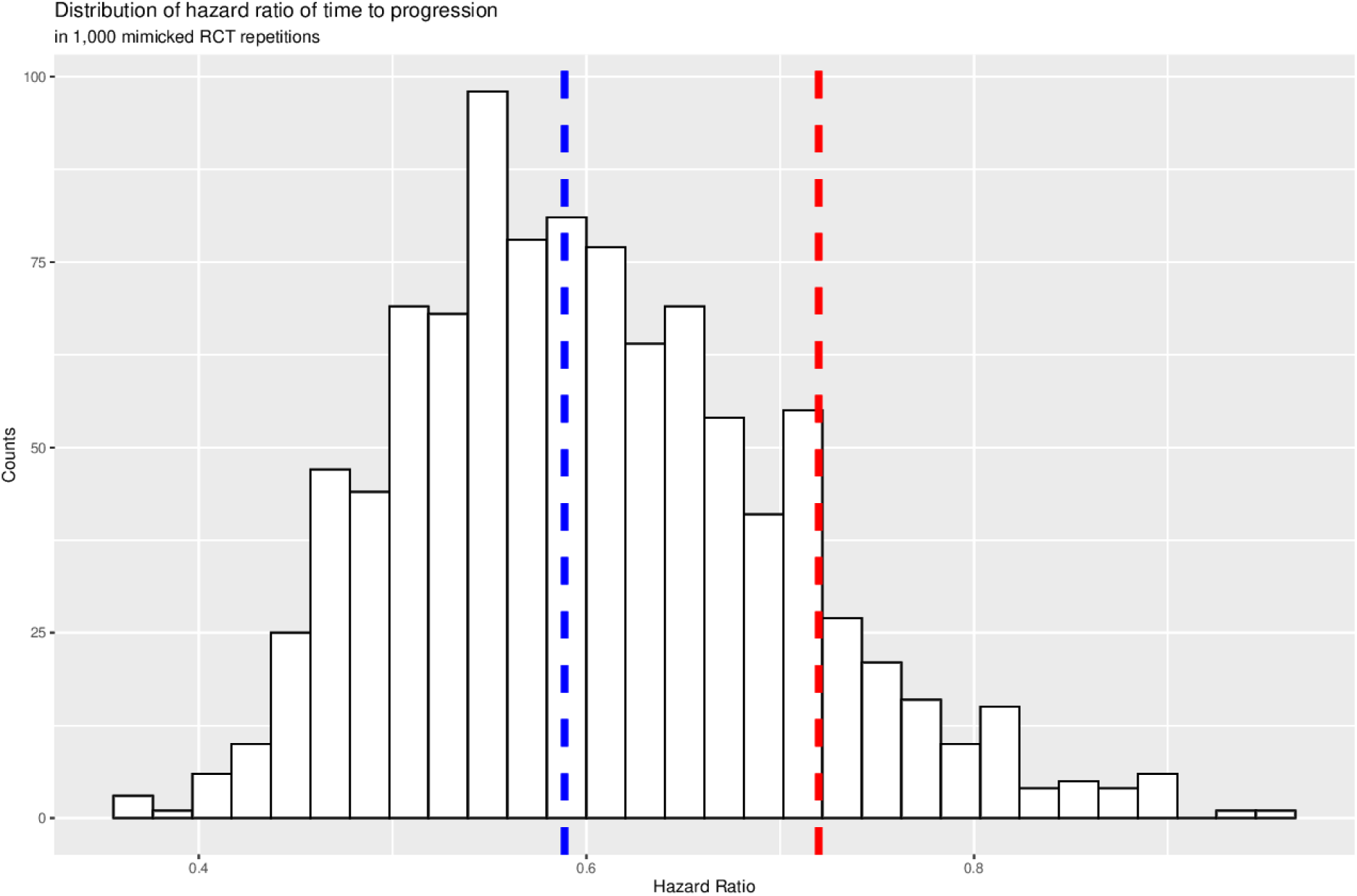
Simulated empirical distributions of A vs B gross efficacy estimates. Distributions of TTP HR obtained over 1,000 replicates of mimicked RCT comparing A to B treatments (N=100 patients per arm) in random samples of the eligible virtual population. Net efficacy estimate obtained with the eligible virtual population (blue dashed line) and gross efficacy of one random mimicked RCT among the 1,000 (red dashed line) are also shown.

The distribution of p-values computed for each of the 1,000 replicates is shown on Figure 3. 10.9% of computed p-values are above 0.05, corresponding to trials showing a non significant HR_gross_.

**Figure 3.**
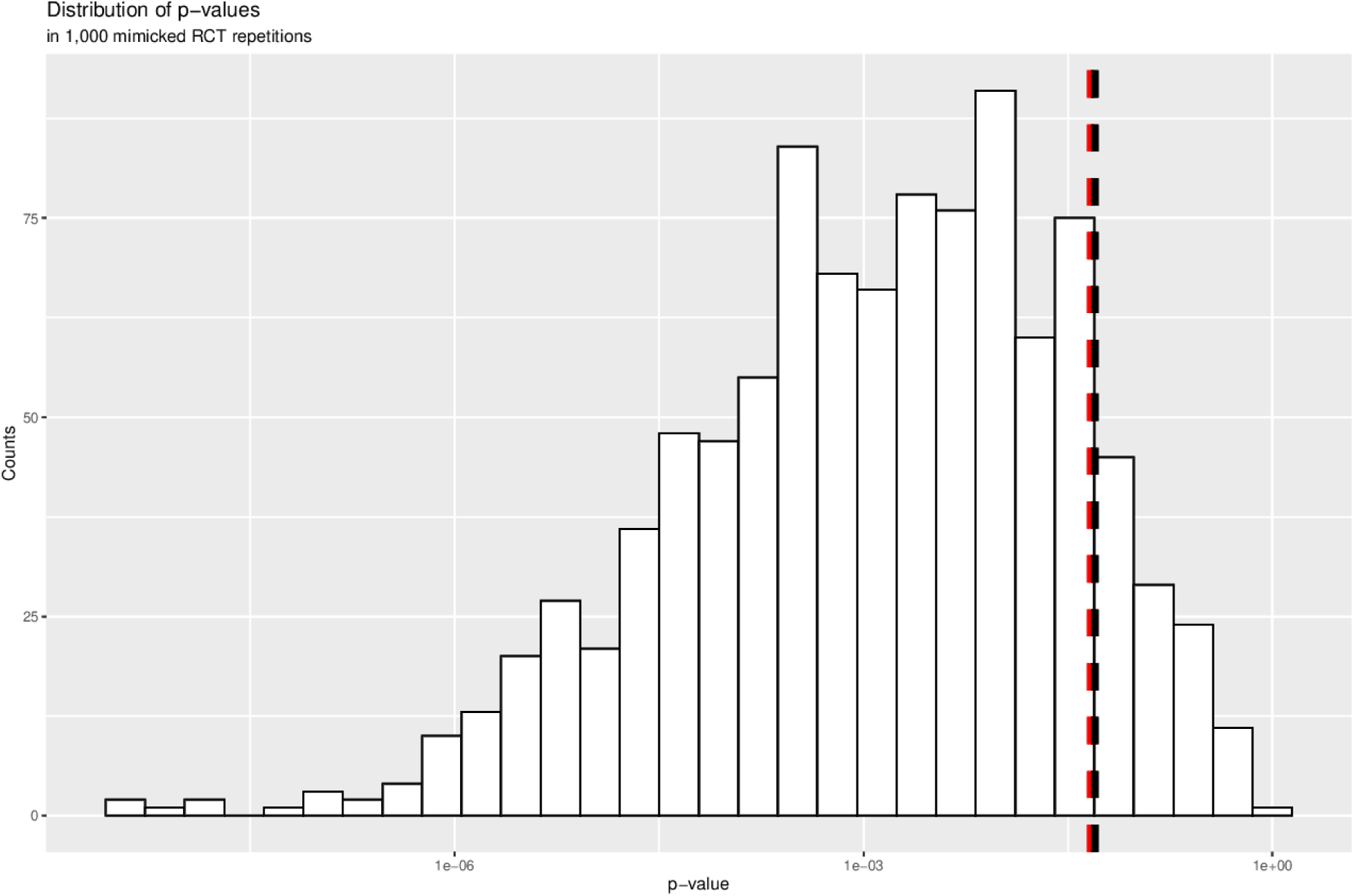
Distribution of p-values extracted from an univariate Cox model. Distributions of p-values obtained over 1,000 replicates of mimicked RCT comparing A to B treatments (N=100 patients per arm) in random samples of the eligible virtual population via a Cox proportional hazards model. The p-value of gross efficacy estimate given by one random mimicked RCT among the 1,000 is also shown (red dashed line). The black dashed vertical line marks the 0.05 statistical significance threshold.

While a real world RCT gives an efficacy estimate that concerns an “average” patient, in silico trials designed to predict NE can in addition provide individual predictions. Figure 4 shows the distribution of individual (net) efficacies on TTP (mean = 3.28 months, sd = 4.45 months) computed among patients who displayed the event of interest (progression) in both treatment arms (n = 1268). These efficacy values are the differences in TTP between B and A. Hence, the above nil values on Figure 4 denote a better TTP with B than with A in the same patients. Out of the 2,000 unique patients, 308 have a tumor that never progresses, 92 have a tumor that progresses under treatment B but not treatment A, and 332 display a tumor progression under treatment A but not under treatment B. The distribution and these figures support that B is more efficacious than A in terms of time to progression.

**Figure 4:**
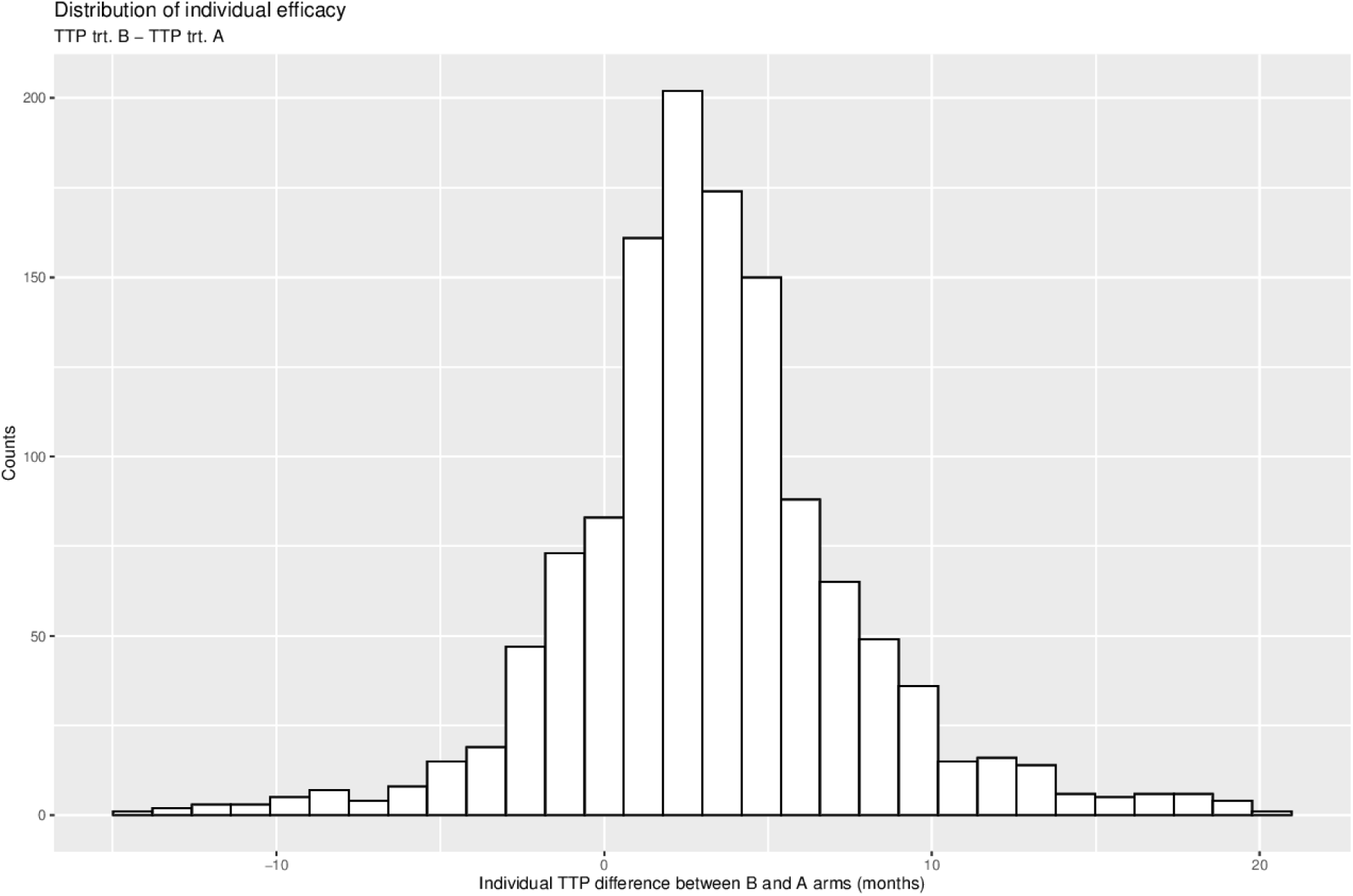
Distribution of individual (net) efficacies on TTP. (mean = 3.28 months, sd = 4.45 months) computed among patients who display the event of interest (progression) in both treatment arms (n = 1268). Out of the 2000 unique patients, 308 have a tumor that never progresses, 92 have a tumor that progresses under treatment B but not treatment A, and 332 display a tumor progression under treatment A but not under treatment B.

In Table 1, the overall results are summarized with the corresponding 95% confidence interval or the PPI. Note that the 95% confidence intervals of GE estimates shown in Table 1 overlap but are not equal: average GE, mimicked RCT that gives the estimate closest to the average GE, random phase 3 ([0.60, 0.61], [0.43; 0.83] and [0.52; 0.99], respectively).

### Mimick of a phase 3 RCT

One RCT among the 1,000 repetitions was randomly chosen to mimic a single phase 3 trial. Its results (the HR using the standard statistical paradigm) are presented in Figure 2 (blue vertical dashed line). Baseline characteristics and demographics of patients randomized in each arm are well matched, as shown in Table 1, allowing the comparison of the two treatment outcomes and the computation of the gross efficacy (HR_gross_) estimate given by this phase 3 trial. In this mimicked phase 3 RCT, treatment A arm had a slightly higher rate of progression at 24 months than did patients in treatment B arm (75% and 73%, respectively; HR_gross_ = 0.72, 95%CI, 0.52 to 0.99, p = 0.046) (Table 1).

## Discussion

### Comments on simulation Findings

These simulations show that modeling and simulation can yield estimates of both net and gross efficacy. Further, they also illustrate the fact that when the estimate of a new treatment efficacy is based on one single RCT, as in a frequent way of doing phase 3 for various more or less compelling reasons, one may well reach efficacy size and/or p-value that wrongly reflect the average gross efficacy or the net efficacy, assumed to be close or even equal to the real efficacy (Figure 2). A single RCT can lead to erroneous conclusions regarding the value of the new treatment (either a non statistically significant difference even if the real efficacy is valuable or an overestimation of its true efficacy). Hence, multiple repetitions of RCTs are required to ensure a reliable estimate of treatment gross efficacy, although this is rarely done or even feasible in real life.

Assuming, i) as a first approximation, a linear multiplicative effect model^27^ for B against A, ii) that the HRs computed in this work are good estimates of the ratio event rate under treatment B over event rate under treatment A, the HR_net_ values correspond for the total population of 2000 patients to the number of progressions prevented by B over A during the 2 years (simulated observation period of 24 months). With the average gross efficacy and the HR_gross_ computed from the mimicked trial closest to the average, the number of prevented progressions are very similar (Table 1). However, with the mimicked phase 3, the number of prevented progressions is approximately 20% less than with HR_net_, corresponding to a markedly reduced population benefit (Table 1). These variations in population benefit estimate of the two treatments are due to differences in patient profiles and thus on responders distributions in the considered trials, everything else alike. They correspond to apparent variation in public health interest of the two treatments.

HR_gross_ obtained over 1,000 simulated mimicked RCTs (gross efficacy estimates) vary from sample to sample (min = 0.35, max = 0.95) because of differences between the two groups due to the randomization process’s inability to generate two groups of patients that are rigorously similar (Figure 2). As expected, the distribution of these HR_gross_ is approximately gaussian. This confirms that two RCTs with the same design and run in the same setting, at the same time, done with two samples drawn randomly from the same population, are likely to have different results and will lead to different population benefit estimates.

The bootstrapped PPI for HR_net_ on progression is [0.52; 0.67], therefore not containing 1, allowing us to conclude that a substantial difference exists between the two treatments. The distribution of the 1,000 observed gross efficacy (HR_gross_) contains the predicted net efficacy (HR_net_) (Figure 2 and Table 1). The figure for net efficacy (HR_net_) is marginally different from the average of the 1,000 mimicked RCTs HR_gross_. However, the numbers of prevented events with treatment B over treatment A are respectively at 240 (net efficacy), 235 (average gross efficacy), 238 (median gross efficacy) and 196 (mimicked phase 3) computed on a base of 2,000 treated patients (Table 1).

Lower than 0.05 in most mimicked RCT, the corresponding p-value for observed gross efficacy varies largely (Figure 3). However, in 10.9% of the mimicked trials, the p-value is larger than 0.05 while all the evidence is in favor of a much more beneficial treatment compared to the other.

The curve of the cumulative mean of HR_gross_ over these 1,000 mimicked trials makes it possible to obtain an insight on the rate of convergence of the repetition of RCTs in this case, i.e. from how many trials the information on efficacy is more or less stable and close to the real efficacy (Figure 1): more than 10 and around 200-250. These numbers are much larger than the 1 or 2 trial repetitions often requested by regulators for the pivotal trial of a new medical product. Simulations as did here suggest that the minimal number depends on the compared treatments, the disease, the selected primary endpoint and the trial targeted population. They also suggest that they can help to find this number.

Would these results be different with other models of another disease and other treatments? Certainly in quantitative terms. Probably not qualitatively since the clues suggested by our findings are based on the gross and net efficacy concepts and RCT methodology rather than specifically on NSCLC and A and B treatments.

## Discussion on the conceptual bases

We will limit the discussion to a few points that we consider fundamental, as a broader discussion would exceed the space allocated for this article.

The idea of comparison to evaluate a therapy while controlling for confounding factors is not new^28^. It led to the first recorded controlled trial. Then came randomization to try to get the most comparable groups of patients possible^29^. Sampling theory and statistical tests were developed at the beginning of the last century. The usefulness of double-blind in minimizing bias became apparent later^30^. Even before the end of the last century, the RCT methodology was mature, while the data analysis was still improving ^31^

What RCT brought to the medical community is the idea that efficacy of a therapy is accessible beyond, or before the fuzzy causal relation between observed patient improvement and the treatment which has been prescribed. The efficacy of a therapy was what the RCT measured. Often, people miss that this measure concerns an average, ideal patient. The concept of net efficacy was born from the possibility offered by modelling and simulation (M&S) to use the subject as his or her own control for simulated trials, even of very long duration, with criteria such as mortality, and large numbers of patients. This is impossible with a real life RCT.

At the beginning of the twenty-first century, the diversification of the type of therapeutics, the raising of new threats to individual and public health, and the deepening of regulatory requirements revealed the limits of the RCT in practice as exemplified recently during Covid-19 pandemic (beyond the lack of international strategy for addressing the curation and prevention challenges)^32^. Costly, time-consuming to set up and complete, and fraught with ethical problems, this methodology does not answer all the questions although it is based on real patients^33^. The possibility of complementing traditional tools with M&S has emerged ^8^. Today, one is exploring this practice and its possibilities^34^. The emergence of the concept of net efficacy is one of these developments. It was thus necessary to better qualify the efficacy measured by an RCT. The first is unbiased, devoid of sources of variability apart from the uncertainty related to its predictive nature (which was not at stake in this work). It is net. The second is subject to variations in the samples, to the inadequacy of the normative nature of randomization. It is gross.

Any model is false and our disease and treatment models cannot deviate from this rule. However, since in our work the same models were used for the calculation of NE and GE and the patients in the trials had the same origin, it follows that comparisons between the HR values found in all trials (NE and GE) are relevant.

We do not observe large differences between the central values of the GE distribution and the predicted NE for the tumor progression linked endpoints. This is reassuring. However, the NE approach has some marked theoretical advantages over gross efficacy, again assuming the model is satisfactorily validated. Beyond the difference in the computational paradigms of NE and GE, the former being totally unbiased while the latter cannot fully control the bias, there is a difference in the populations of interest. The population concerned by NE is the entire population at risk if the modeler has been able to construct a sufficiently realistic virtual population. With the GE in real-life randomized trial, the relevant population must be inferred from the sample on which this GE is calculated. It follows that with real-life RCT, the parent population of the sample on which the treatment is tested is, at best, only a portion of the actual population of interest^35^. This population of interest is represented by the virtual population in the case of the NE calculation. Further to being guaranteed without bias, the NE prediction is obtained at no ethical burden, at a much lower cost and more rapidly than even a single GE estimate.

Special cases include cross-over designed trials and N-of-1 trials, which allow within-patient comparisons. The former results in an average GE, as a parallel group design. The latter, as for NE, results in an individual efficacy estimate. However, both designs are dependent on time.

And last but not the least, thanks to the way it is computed, NE opens to responder profiling and personalized medicine^36^. We gather evidence on therapy efficacy with the ultimate goal of deciding whether this therapy is good for our patient^37^. While the analysis of RCT data that allows to estimate GE even cannot guarantee sub-group findings, limitation which, associated with restrictions in sampled minorities^38^ impairs the derivation of personalizing therapy algorithms. And, with individual NE value, the NE computation provides a prediction of benefit due to the new treatment over its comparator for each of the patients of the virtual population while any single estimate of GE concerns a single average patient. Note that this average patient is virtual.

This remark leads to the following fundamental question: what are we trying to measure with GE or NE efficacy? In both cases, it is an average quantity and therefore virtual because it does not concern any real patient. The GE efficacy can only remain at the group level because this is how it can be calculated with the minimum residual bias while the NE efficacy can go down to the level of the individual patient since it is the sum of the individual NEs that the M&S approach allows to predict (Figure 4). What is "real" efficacy? It is neither gross nor net efficacy, even if, because of its construction with the subject, its own witness, the latter should be close to it. This question goes back to the question of the nature of therapeutic efficacy. This is not the place to open this epistemological debate. One element of the debate seems to us to be provided by a practical difference between net efficacy and gross efficacy. With respect to the HR chosen as the efficacy metric in our illustration, it can be predicted individually in the net efficacy calculation, whereas it is calculated over the entire sample in the gross efficacy calculation. It is clear from this case that a distinction must be made in reality between individual efficacy and average efficacy. It is tempting to define a therapy efficacy as the anticipated change in disease course because of a prescription.

The phase 3 trial randomly selected among the 1,000 mimicked RCTs led to a statistically significant efficacy. However, this efficacy is different in quantity both from that predicted by the NE and from the average of the 1,000 HR GE efficacy distribution. These HRs lead to different NPEs which would be reflected in different assessed treatment benefits (Table 1). Further, the distribution shown in Figure 2 shows that if most trials result in GE close to NE, it remains possible that a single trial gives a rather different estimate of efficacy.

Our results suggest that under certain circumstances, while NE may indicate only a marginal benefit, consistent with an unfavorable benefit-risk ratio, a real-life clinical trial could be statistically significant. Decision-makers who emphasize the statistical significance without considering efficacy size may erroneously judge a treatment to be beneficial.

Table 2 shows that the subpopulations allocated to the treatment arms of this trial are not different and do not differ from the overall population for the variables measured in real life characterizing the patients. However, our results show that there are differences in the values of calculated efficacy and the numbers of prevented events. One possible explanation for this discrepancy is the impact on patients’ treatment effect of variables that are not measured in real life (the vast majority of patient descriptors in our case), and as such cannot impact on the arms baseline characteristic comparison, but are taken into account in the models and thus in its predictions.

**Table 2.**
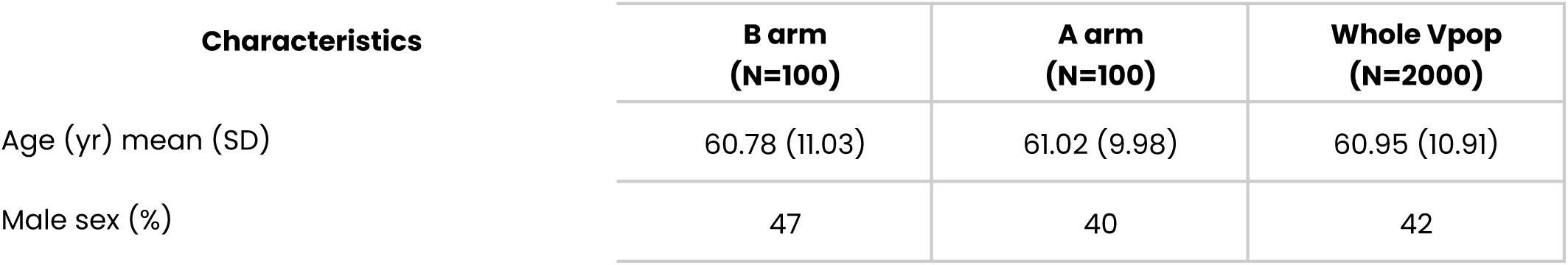
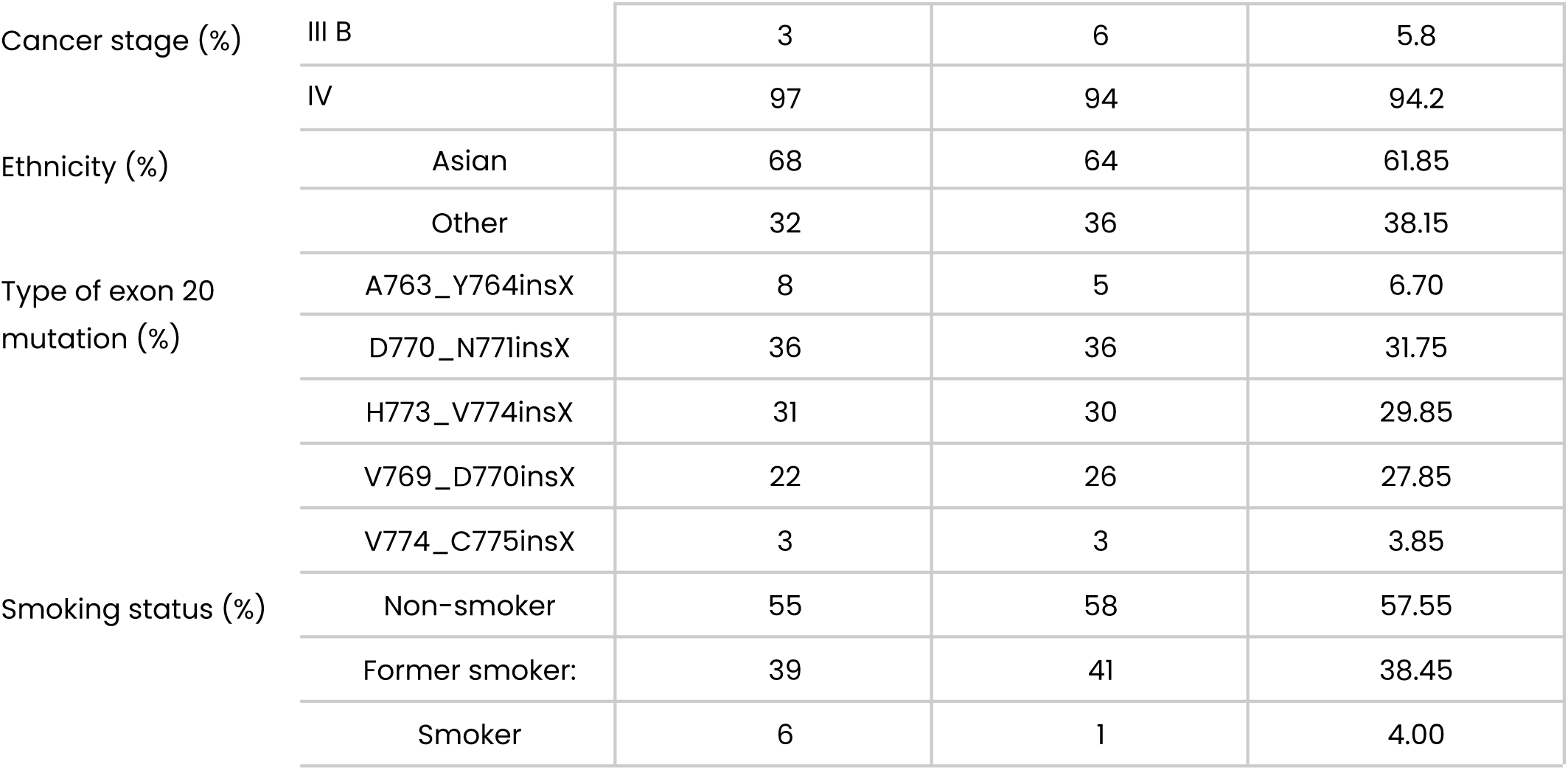
Baseline characteristics and demographics of virtual patients allocated to A and B arms in the phase 3 mimicked RCT.

| Characteristics | B arm<br>(N=100) | A arm<br>(N=100) | Whole Vpop<br>(N=2000) |
| --- | --- | --- | --- |
| Age (yr) mean (SD) | 60.78 (11.03) | 61.02 (9.98) | 60.95 (10.91) |
| Male sex (%) | 47 | 40 | 42 |

|  |  |  |  |  |
| --- | --- | --- | --- | --- |
| Cancer stage (%) | III B | 3 | 6 | 5.8 |
|  | IV | 97 | 94 | 94.2 |
| Ethnicity (%) | Asian | 68 | 64 | 61.85 |
|  | Other | 32 | 36 | 38.15 |
| Type of exon 20 mutation (%) | A763_Y764insX | 8 | 5 | 6.70 |
|  | D770_N771insX | 36 | 36 | 31.75 |
|  | H773_V774insX | 31 | 30 | 29.85 |
|  | V769_D770insX | 22 | 26 | 27.85 |
|  | V774_C775insX | 3 | 3 | 3.85 |
| Smoking status (%) | Non-smoker | 55 | 58 | 57.55 |
|  | Former smoker: | 39 | 41 | 38.45 |
|  | Smoker | 6 | 1 | 4.00 |

From these results, it can be said that, whether generally QSP can duplicate real life RCT results and help in exploring response factors^39^, the GE approach can answer the question of the causal relationship between the tested treatment and its expected effect, while the NE approach, in addition, and beyond to establish the mechanical background of this effect, leverages a prediction at the population or even individual level of the amount of efficacy of the treatment.

As mentioned above, the disease model, the treatment models and the virtual population used in the illustration of the two concepts have been validated. Model or virtual population validation can never be universal and definitive. It can only be evaluated, based on various considerations, as sufficiency in relation to the objective of the modelling and the context of use^40^. However, whatever is the degree of validity of the models - which is deemed good in this example, especially because the models were able to predict prospectively and blindly the outcomes of a real RCT - it does not influence the qualitative results presented here because of the comparative approach.

The possible values of net efficacy are a continuum. There is no natural threshold of values between sufficient and unnecessary treatments. The HR value of 1, which reflects absolute inefficacy, limits the range of values corresponding to a lower efficacy than the control. Beyond 1, the new treatment is less effective compared to the control. Below 1, it is more effective but there is no threshold where it becomes more beneficial in terms of public or individual health. The concept of net efficacy does not incorporate a process that translates an insufficient amount of efficacy into the non-efficacy of the new treatment over the control.

On the contrary, with the gross efficacy this translation is provided by the statistical test that can be integrated into its construction. It helps to distinguish between the treatment that can be considered efficacious (significant unilateral testing) and the one that can or should be rejected. This is where it is necessary to distinguish between the calculation of the size of the efficacy of the treatment being tested and the interpretation of this size, which determines its “efficacy”.

With net efficacy, you have to imagine a predefined threshold for defining an impactful size of efficacy. Note that this threshold exists in the case of gross efficacy. It is implicitly introduced in the calculation of the number of subjects to be included in the RCT. The discussion of the threshold is outside the scope of this article^41^.

The virtual population is a key component in the calculation of net efficacy. It should represent the population of interest as realistically as possible^21^. This will need to be ensured by applying a recognized validation process^25^. In the case where each virtual patient is a replica - or a twin - of a real patient, the virtual population is a population of digital twins^42^.

The mimicked RCT in our illustration does not exactly mimic the actual RCT. This is because, in real life, the sample that is the focus of the trial does not come directly from the eligible population of interest, since patients are already selected because the sites participating in the trial are not a random sample of the various health care organizations through which patients from the population of interest access the health care system. However, this may not be the case for some rare diseases that are centrally managed. This, in addition to the not always rigorous application of eligibility criteria, explains why the trial sample is not randomly drawn from the population of interest. However, we assumed that the samples of the mimicked RCT were random. So, in reality, the differences between the values of gross efficacy and net efficacy could be larger than those in our example.

## Conclusion

Net efficacy represents a novel conceptualization of therapeutic efficacy. Predicted through modeling and simulation, it is unbiased by design and individualized, likely approximating true efficacy more closely than gross efficacy measured by RCTs.

In these simulations, the ratio of the sample size of the RCTs to the population of interest size is rather high. With a lower ratio, corresponding to smaller samples or a larger population, the results would be quantitatively different but qualitatively identical. While the HRs calculated by the GE approach and by the NE approach are not very different, they could nevertheless lead to rather different assessments of the benefit of the new treatment compared to its comparator.

This work also shows that a phase 3 trial of a truly effective product can very well be negative by current standards and therefore wrongly lead to its withdrawal. Carrying out a single RCT in phase 3 is therefore taking an industrial and ethical risk. But it is impossible to include in the development plan a sufficient number of trials that would guarantee a risk-free decision. If a single phase 3 trial is not enough, enough trials are impractical.

But the prediction of net efficacy can serve as a guide for the development of a new therapy. It can first be used to compute the sample size of RCTs. It can also be used as a reference if the Phase 3 RCT is negative or inconclusive and can correct the interpretation of trials that are false-negative or have led to efficacy estimates that are deemed too small or of less-than-acceptable strength of evidence (because, e.g., a small number of events despite a statistically significant difference). With an unquestionable phase 3 trial, computation of the NE will enable a more relevant estimate of the population benefit. It can also serve as a starting point for a Bayesian process that would increase the chances of a reliable conclusion for an acceptable total cost. Another option would be to consider the prediction of the net efficacy made at the end of phase 2 as reality, with only one, or even two underpowered RCTs or even a small cohort in phase 3 which would aim at an ultimate validation of the models, validation that would complete the credibility of the models and therefore their predictions obtained on a validated representation of the population of interest, allowing the model predictions to complete the information obtained from clinical trials that remain the core of the new treatment assessment. And, last but not the least the M&S computation of the NE can help to solve the ethical dilemma pointed out by Milton Packer when replicating a well designed RCT that has shown survival benefit in heart failure^43^.

#### Take home messages

- The notion of therapeutic efficacy is difficult to define; in silico, it is possible to approach the two types of efficacy that can be conceptualized, net efficacy (NE) and gross efficacy (GE), the one accessible to RCTs
- Variability in RCT results for a given actual efficacy may lead to a trial negative result even when the treatment is more effective than its control
- And the reverse is also possible
- In silico prediction of net efficacy (NE) can help understanding a situation created by the uncertain results of a pivotal Phase 3 trial
- This leads to the idea of a clinical development in which clinical studies and the in silico approach would be combined for faster, less expensive and more ethical development
- While the RCT gives an estimate of efficacy on an ideal average patient, the in silico clinical trial (ISCT) allows for individualized prediction of efficacy
- The statistical and the in silico (here its so-called physiological level) paradigms are not in competition; they are complementary if not intertwined
- The in silico paradigm embedded a large array of tools: PK, PBPK, QSP (including the knowledge-based modelling approach used here), AI
- For regulatory decision making, we cannot imagine NE alone, without any real-life based evidence, i.e. GE estimate(s) from real data, at least for validating the computed NE.

## Data Availability

The datasets generated and/or analyzed during the current study are available from the corresponding author on reasonable request. The code utilized during the current study can be made available on the jinkō.ai platform by the corresponding author on reasonable request.

https://jinko.ai/

## Acknowledgements

The authors would like to thank Jean-Christophe Lega for his review and insightful comments and suggestions.

## Author contributions

J.-P. B., R. K., C. M., E. P., and M. H. conceived the project and designed the study. J.-P.B, E.B. and S.G.N. worked on definitions and concepts. A. N., C. M., and M. D. developed the model. E. J., R. K., J.-P. B., and E. P. handled methodology, statistics, and visualization. J.-P. B., M. D., E. P., M. H., and C. M. performed clinical validation and interpretation. All authors drafted the manuscript, provided critical review and approved the final manuscript.

## Ethics

This research involved only computationally generated virtual patients and did not include any human participants, human material, or human data. Therefore, ethics committee approval and informed consent were not required.

## Supplementary information

N/A

